# Calprotectin and inflammation-associated serum biomarkers determine critical illness in COVID-19

**DOI:** 10.1101/2022.01.28.22270001

**Authors:** Georgios Kassianides, Athanassios Siampanos, Garyphalia Poulakou, George Adamis, Aggeliki Rapti, Haralampos Milionis, George N. Dalekos, Vasileios Petrakis, Styliani Sympardi, Symeon Metallidis, Zoi Alexiou, Theologia Gkavogianni, Evangelos J. Giamarellos-Bourboulis, Theoharis C. Theoharides

**Author notes:** **Corresponding author:** E. J. Giamarellos-Bourboulis, MD, PhD, 4^th^ Department of Internal Medicine, ATTIKON University Hospital, 1 Rimini Street; 124 62 Athens; Greece. equal contribution (Evangelos J. Giamarellos-Bourboulis, Theoharis C. Theoharides).

## Abstract

Little is known on the key contributing factors towards progression into acute respiratory distress syndrome (ARDS) necessitating mechanical ventilation (MV) in COVID-19. We determined serum levels, within 24 hours of diagnosis, of alarmins, as well as pro- and anti-inflammatory molecules in asymptomatic, moderate, severe and intubated patients compared to non-infected comparators. Levels of the pro-inflammatory interleukin (IL)-8, IL-18, matrix metalloproteinase-9, platelet-derived growth factor (PDGF)-B and calprotectin (S100A8/A9) were specific drivers of ARDS. Levels of the anti-inflammatory IL-1ra and IL-33r were increased; IL-38 was increased only in asymptomatic patients, but significantly decreased in the more severe COVID-19 cases. Multivariate ordinal regression showed that pathways of IL-6, IL-33 and calprotectin gave significant probability for worse outcome. These results indicate a dysfunctional response to the presence of alarmins that may be used for prognosis and development of effective treatments.

---

The SARS-CoV-2 coronavirus infects human cells by first binding to their surface receptor, Angiotensin Converting Enzyme 2 (ACE2), via its corona spike protein^1^ leading to the development of COVID-19^2,3^. This condition involves a complex immune response that includes the release of a “storm” of pro-inflammatory cytokines^4-6^ associated with poor prognosis. Prominent among them have been IL-6^7-9^ and IL-1β^10^ but also the family of S100 alarmins, the S100A8/A9 of which is otherwise known as calprotectin^11^.

This knowledge led early to consider severe COVID-19 as a hyper-inflammatory disorder and encouraged the administration of modifiers of the biological response like dexamethasone, tocilizumab, anakinra and baricitinib in the forefront of management. However, after almost two years in this pandemic, we have realized that when severe COVID-19 is further progressing into acute respiratory distress syndrome (ARDS) necessitating mechanical ventilation (MV) the prognosis is poor. This fact emphasizes the need to identify the real drivers leading from moderate and severe COVID-19 to severe ARDS and MV. This may be the only way to prevent or efficiently address the most ominous consequence of COVID-19.

In this paper, we investigated serum levels of alarmins, as well as pro- and anti-inflammatory molecules leading to ARDS and MV, and how their presence tracks with disease severity.

## RESULTS

### Study participants

The sampling for the study took place between April and November 2020. Serum samples from 181 patients and 40 non-infected comparators were studied. Among the patient population, on the day of blood sampling 19 patients were asymptomatic; 42 patients had moderate disease; 78 patients had severe disease; and 42 patients were with ARDS on MV. Patients with ARDS on MV had higher values of the sequential organ failure assessment (SOFA) score, lower values of respiratory ratios and higher circulating C-reactive protein and ferritin (Table 1).

**Table 1.** Demographic characteristics of participants at the first stage of the study.

|  | **Comparators** | **Asymptomatic** | **Moderate** | **Severe** | **ARDS and MV** |
| --- | --- | --- | --- | --- | --- |
| Number | 40 | 19 | 42 | 78 | 42 |
| Age (years) mean ± SD | 58.3 ± 15.8 | 59.5 ± 10.8 | 55.3 ± 14.8 | 61.4 ± 13.9 | 64.9 ± 12.8 |
| Male gender, n (%) | 28 (70) | 12 (63.2) | 25 (59.5) | 56 (71.8) | 35 (83.3) |
| SOFA, mean ± SD | NA | NA | 0.9 ± 0.9 | 2.3 ± 1.6 | 6.5 ± 2.5 |
| CCI, mean ± SD | 1.0 ± 0.2 | 1.3 ± 1.9 | 2.3 ± 2.3 | 2.6 ± 2.1 | 2.4 ± 1.5 |
| Comorbidities, n (%) |  |  |  |  |  |
| Type 2 diabetes mellitus | 0 (0) | 5 (26.3) | 13 (31.0) | 20 (25.6) | 6 (14.3) |
| Chronic heart failure | 0 (0) | 0 (0) | 3 (7.1) | 5 (5.1) | 0 (0) |
| Chronic renal disease | 0 (0) | 0 (0) | 2 (4.8) | 3 (3.8) | 2 (4.8) |
| Coronary heart disease | 0 (0) | 0 (0) | 5 (11.9) | 8 (10.3) | 6 (14.3) |
| Dyslipidemia | 0 (0) | 4 (21.1) | 14 (33.3) | 17 (21.8) | 4 (9.5) |
| Hypothyroidism | 0 (0) | 2 (10.5) | 7 (16.7) | 7 (9.0) | 3 (7.1) |
| Hypertension | 0 (0) | 2 (10.5) | 16 (38.1) | 24 (30.8) | 7 (16.7) |
| Stroke | 0 (0) | 0 (0) | 2 (4.8) | 1 (1.3) | 1 (2.4) |
| Atrial fibrillation | 0 (0) | 0 (0) | 2 (4.8) | 6 (7.7) | 3 (7.1) |
| COPD | 0 (0) | 0 (0) | 3 (7.1) | 2 (2.6) | 0 (0) |
| pO<sub>2</sub>/FiO<sub>2</sub>, mean ± SD | NA | NA | 397.5 ± 84.3 | 274.5 ± 103.8 | 127.1 ± 72.2 |
| White blood cells count, mean ± SD (/mm<sup>3</sup>) | NA | NA | 5394.3 ± 2379.8 | 7000.1 ± 2945.8 | 11977.8 ± 6094.2 |
| CRP, median (IQR), mg/l | NA | NA | 4.9 (30.6) | 39.0 (80.1) | 82.9 (173.5) |
| Ferritin, median (IQR),<br>ng/ml | NA | NA | 293.2<br>(534.2) | 485.3<br>(832.5) | 1189.5 (1686.8) |

Abbreviations ARDS= acute respiratory distress syndrome; CCI= Charlson's comorbidity
index; CRP= C-reactive protein; COPD= chronic obstructive pulmonary disease; FiO<sub>2</sub>:
fraction of inspired oxygen; IQR= interquartile range; MV= mechanical ventilation;
NA=not available; pO<sub>2</sub>: partial oxygen pressure; SD= standard deviation; SOFA
sequential organ failure assessment score

At the first stage of the study, the samples were analyzed in an effort to identify a set of mediators that may drive progression into ARDS and MV. At the second stage of the study, the mediators were serially followed over-time to define if the levels change over-time until ARDS and MV (Figure 1).

**Figure 1.**
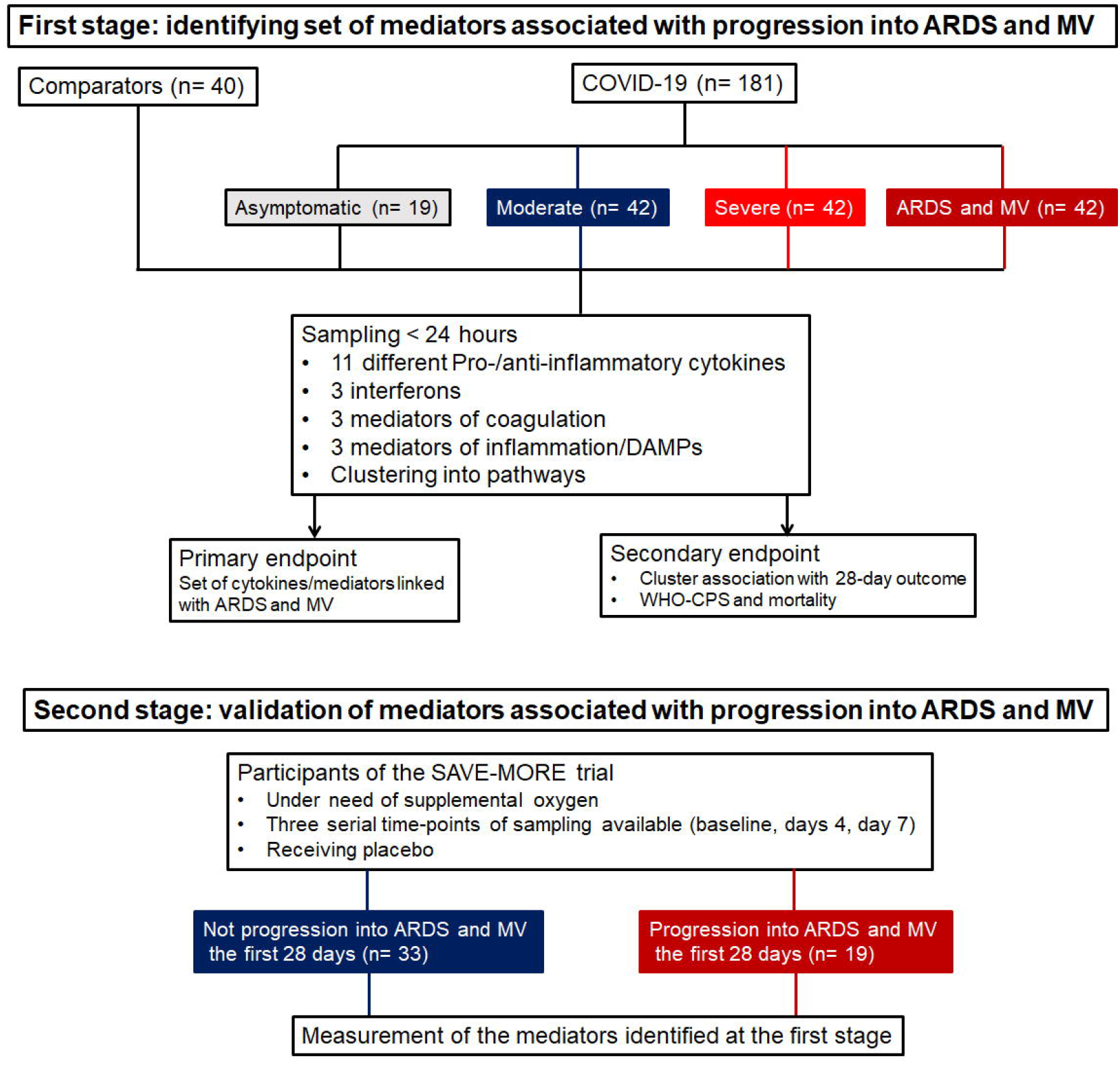
Study flow chart. Abbreviations: ARDS= acute respiratory distress syndrome; MV= mechanical ventilation; n=number of patients

### First stage: Presence of inflammation-associated mediators

Among the pro-inflammatory mediators, interleukin (IL)-1β, IL-6, IL-17, IL-33 and tumour necrosis factor-alpha (TNFα) did not differ between the different stages of severity. However, IL-8 and IL-18 were significantly greater among patients with critical ARDS necessitating MV compared to less severely ill patients. Among anti-inflammatory cytokines, IL-33r (souble ST2) was significantly greater among patients at critical ARDS. IL-1ra (soluble receptor antagonist) was greater among patients with mild to critical COVID-19 without, however, significant difference between those with severe and critical disease. IL-10 did not differ among patients and controls. IL-38 levels were decreased from asymptomatic patients to the more severe stages (Figure 2). Interferons did not differ between patients (Supplementary Figure 1).

**Figure 2.**
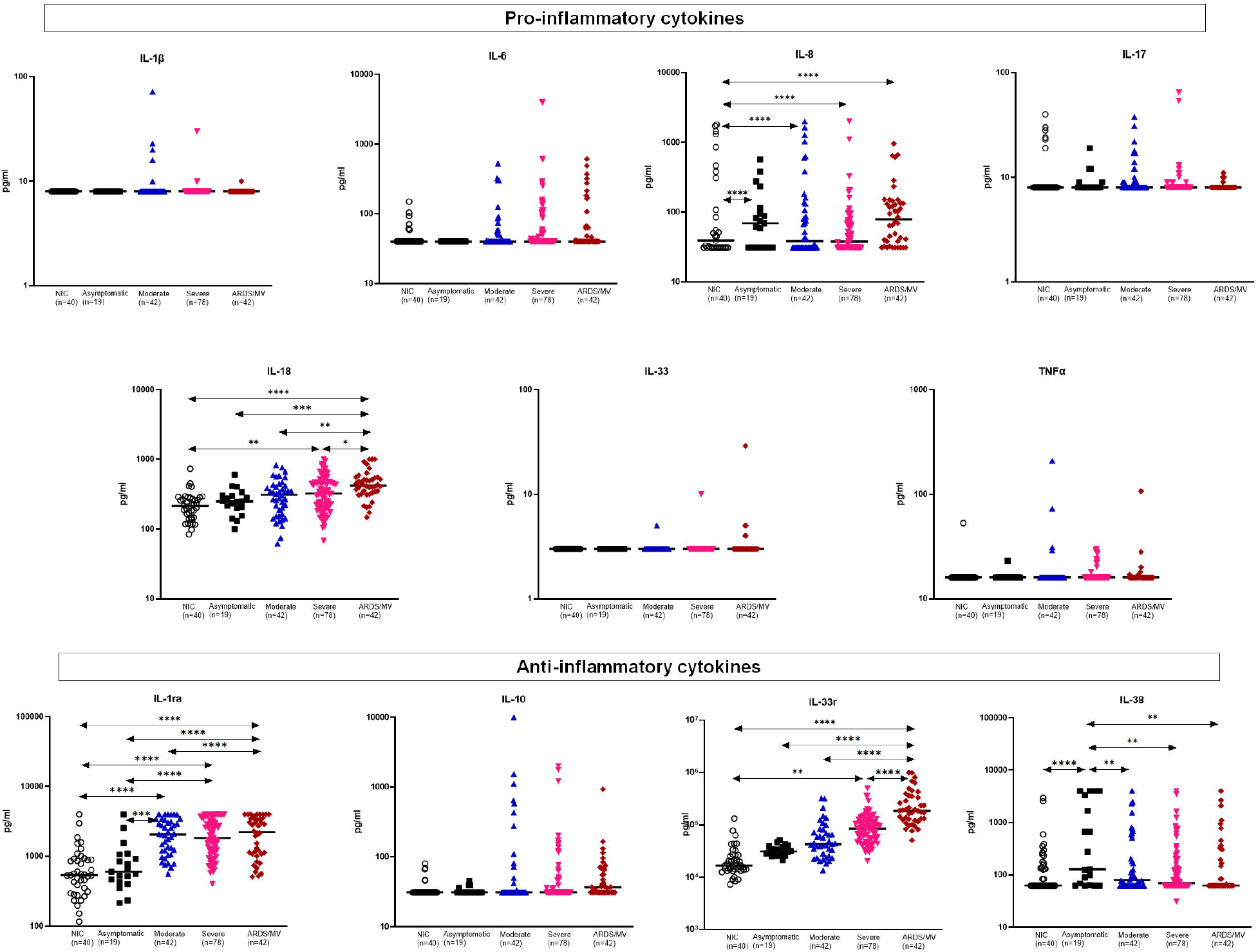
Concentrations of pro-inflammatory and anti-inflammatory cytokines. Dot plots with horizontal lines indicating the median of each group. The numbers of subjects evaluated are listed in the parentheses. Double arrowhead lines indicate comparisons between groups. Only statistically significant differences are indicated as follows: *p <0.05; ** p <0.01; ***p< 0.001; ****p< 0.0001. Abbreviations: ARDS= acute respiratory distress syndrome; IL=interleukin; MV= mechanical ventilation; n=number of patients; NIC= non-infected comparators; TNFα=tumor necrosis factor-alpha.

Platelet-derived growth factor (PDGF)-B, and to a lesser extent PDGF-A, were increased among critically ill ARDS; PAF did not differ. The same was the case for calprotectin and matrix metalloproteinase (MMP)-9. PAF and S100B did not differ among different stages of severity (Figure 3).

**Figure 3.**
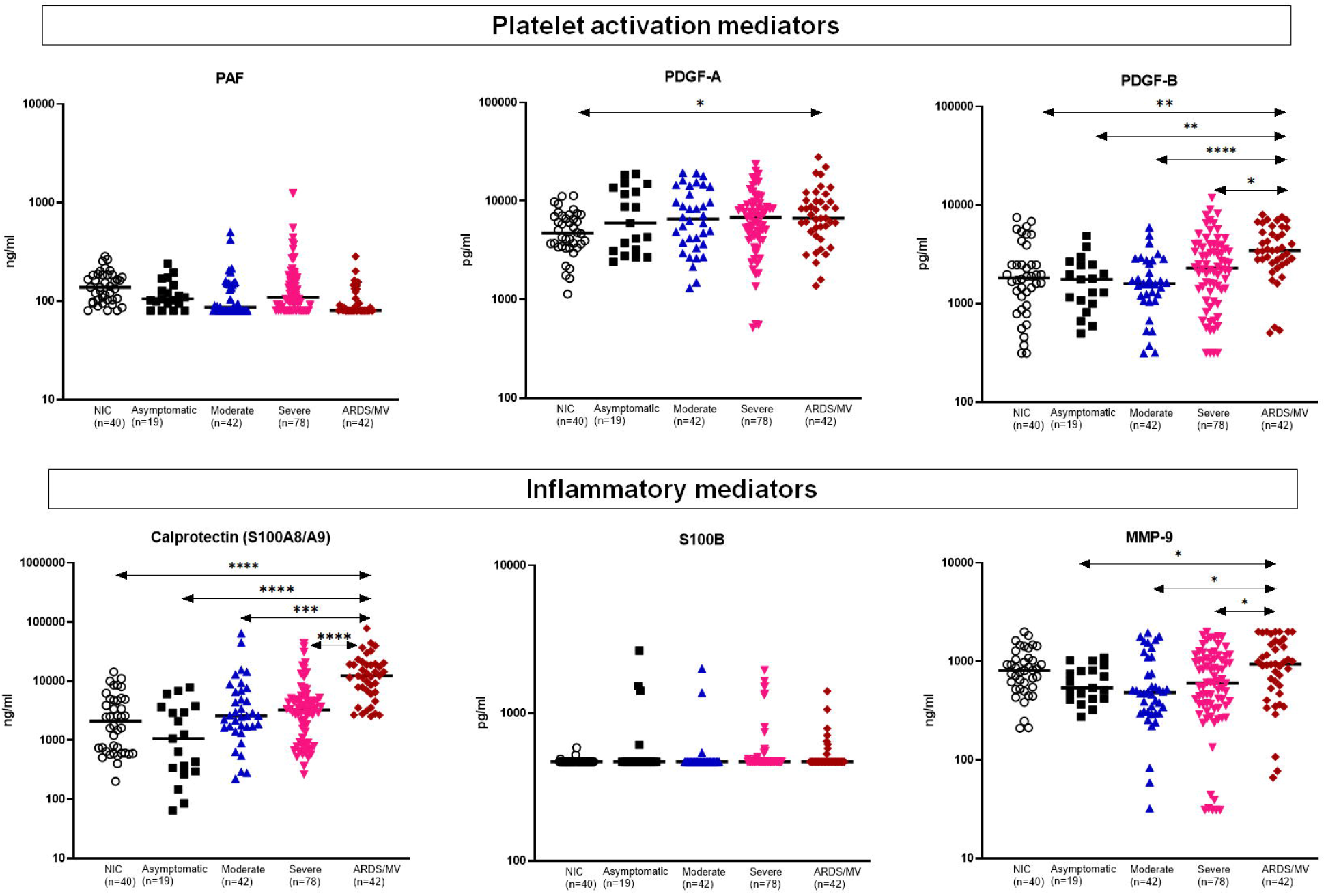
Concentrations of platelet-associated mediators and of other measured inflammatory mediators. Dot plots with horizontal lines indicating the median of each group. The numbers of subjects evaluated are listed in the parentheses. Double arrowhead lines indicate comparisons between groups. Only statistically significant differences are indicated as follows: *p <0.05; ** p <0.01; ***p< 0.001; ****p< 0.0001. Abbreviations: ARDS= acute respiratory distress syndrome; MMP: matrix metallo-proteinase; MV= mechanical ventilation; n=number of patients; NIC= non-infected comparators; PAF=platelet activation factor; PDG= platelet derived growth factor

### First stage: drivers of acute respiratory distress syndrome (ARDS) necessitating mechanical ventilation (MV)

ROC curve analyses identified that IL-8, IL-18, MMP-9, IL-33r, PDGF-B and calprotectin were associated with ARDS necessitating MV (Figure 4A). The cut-offs of each of the six mediators providing the best trade-off of sensitivity and specificity were defined and entered the equation of multivariate analysis. Only increased IL-33r, increased PDGF-B and increased calprotectin were found to be drivers of ARDS necessitating MV (Figure 4B).

**Figure 4.**
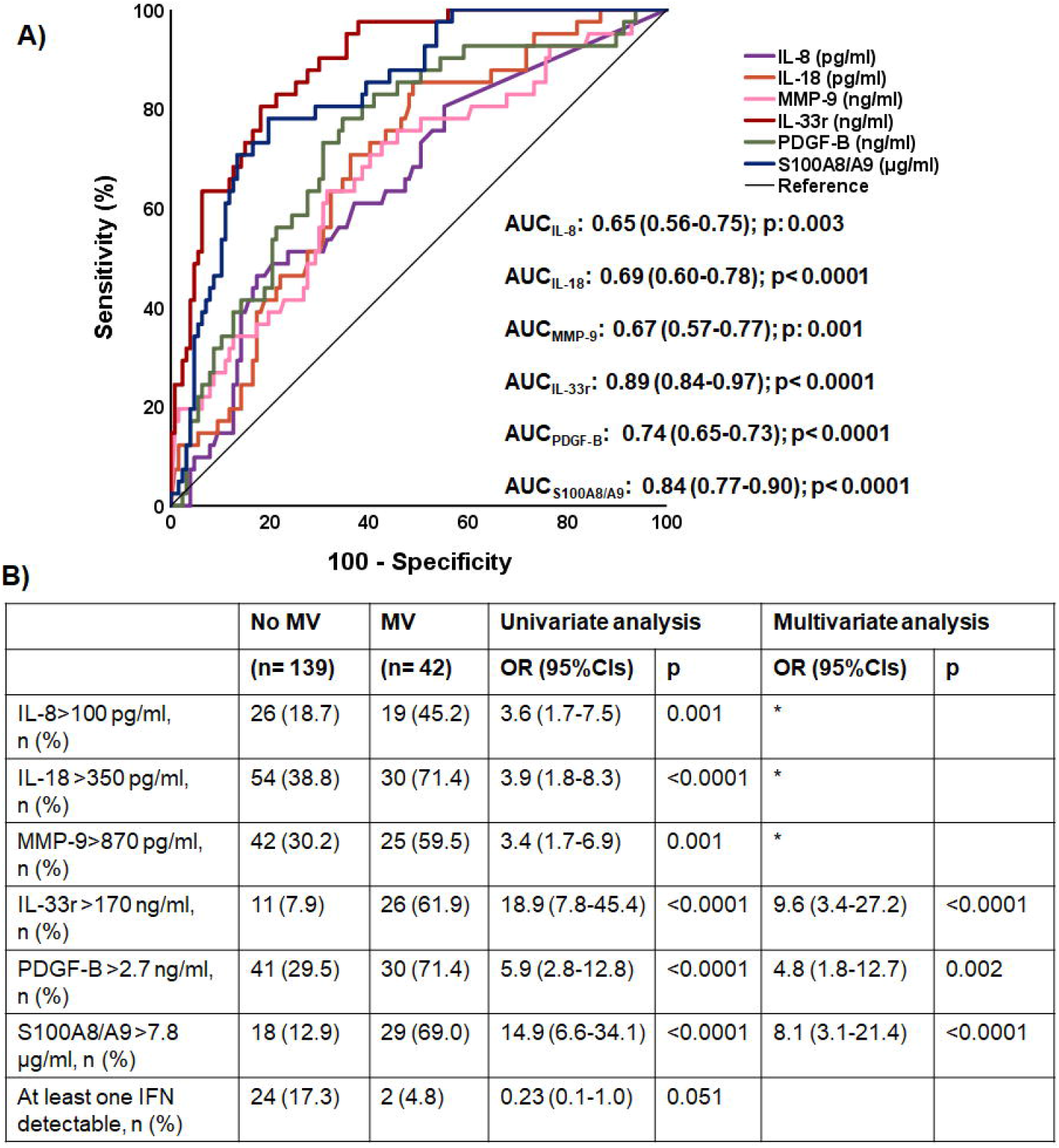
Main drivers of acute respiratory distress syndrome (ARDS) necessitating mechanical ventilation. (A). Receiver operator characteristics (ROC) curves of the six studied mediators that are providing statistically significant area under the ROC curve (AUC) for the detection of ARDS necessitating mechanical ventilation (MV). The AUC, their 95% confidence intervals and the p-values of significance are provided. The ROC curves of the other 14 mediators measured are not provided because they did not have any statistical significance. (B). Following Youden index analysis, the cut-offs of each of the six mediators shown in panel A were determined. These cut-offs entered univariate and multivariate forward stepwise logistic regression analyses to select the mediators associated with ARDS necessitating MV. *variables excluded after 3 steps of forward analysis. Abbreviations: CI=confidence interval; OR=odds ratio.

### First stage: drivers of 28-day mortality

The analysis involved hospitalized patients with moderate and severe COVID-19 and patients with ARDS necessitating MV. For this analysis, we decided to follow a pathway-like division of biomarkers into clusters. To this end, nine pathways were defined as follows: a) IL-1ra activation when both IL-1β and IL-1ra were above the median of the entire cohort; b) IL-6 pathway when IL-6 was above the median of the entire cohort; c) IL-18 pathway when IL-18 was above the cut-off defined for critical illness; d) neutrophil activation when at least two of IL-8 above >100 pg/ml, MMP-9 above 870 pg/ml or IL-17 above the lower limit of detection were met; e) interferon pathway when at least one of the three measured interferons was above the median of the entire cohort; f) IL-33 pathway when IL-33r was above 170 ng/ml; g) IL-38 pathway when IL-38 was above the median of the entire cohort; h) PDGF-B pathway when PDGF-B was above 2.7 ng/ml; and i) calprotectin pathway when calprotectin was above 7.8 μg/ml (Figure 5 and Figure 6A).

**Figure 5.**
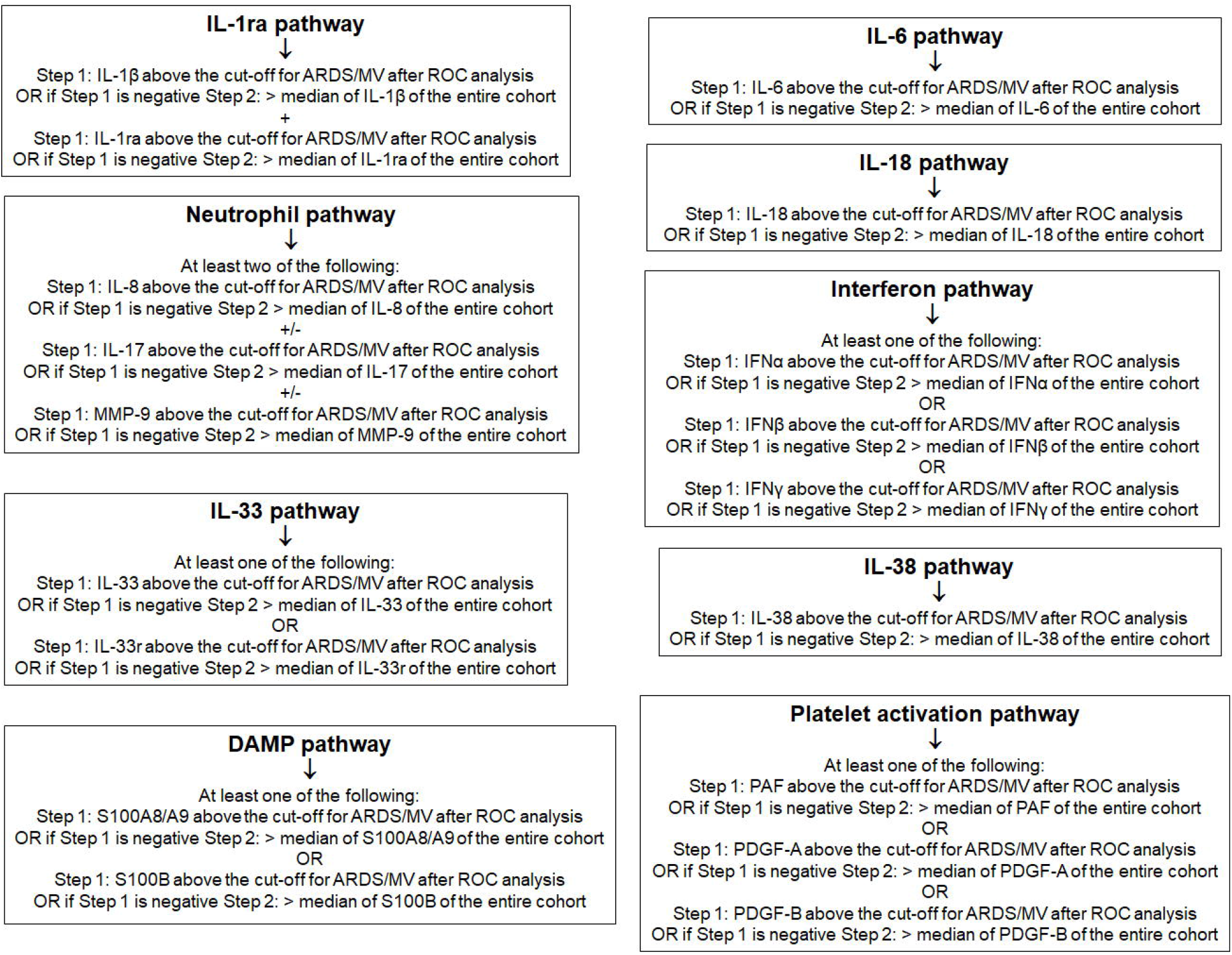
Principles of pathway analysis. Abbreviations: ARDS= acute respiratory distress syndrome; DAMP= danger-associated molecular pattern; IFN= interferon; IL= interleukin; MV= mechanical ventilation; PAF=platelet activation factor; PDG= platelet derived growth factor

**Figure 6.**
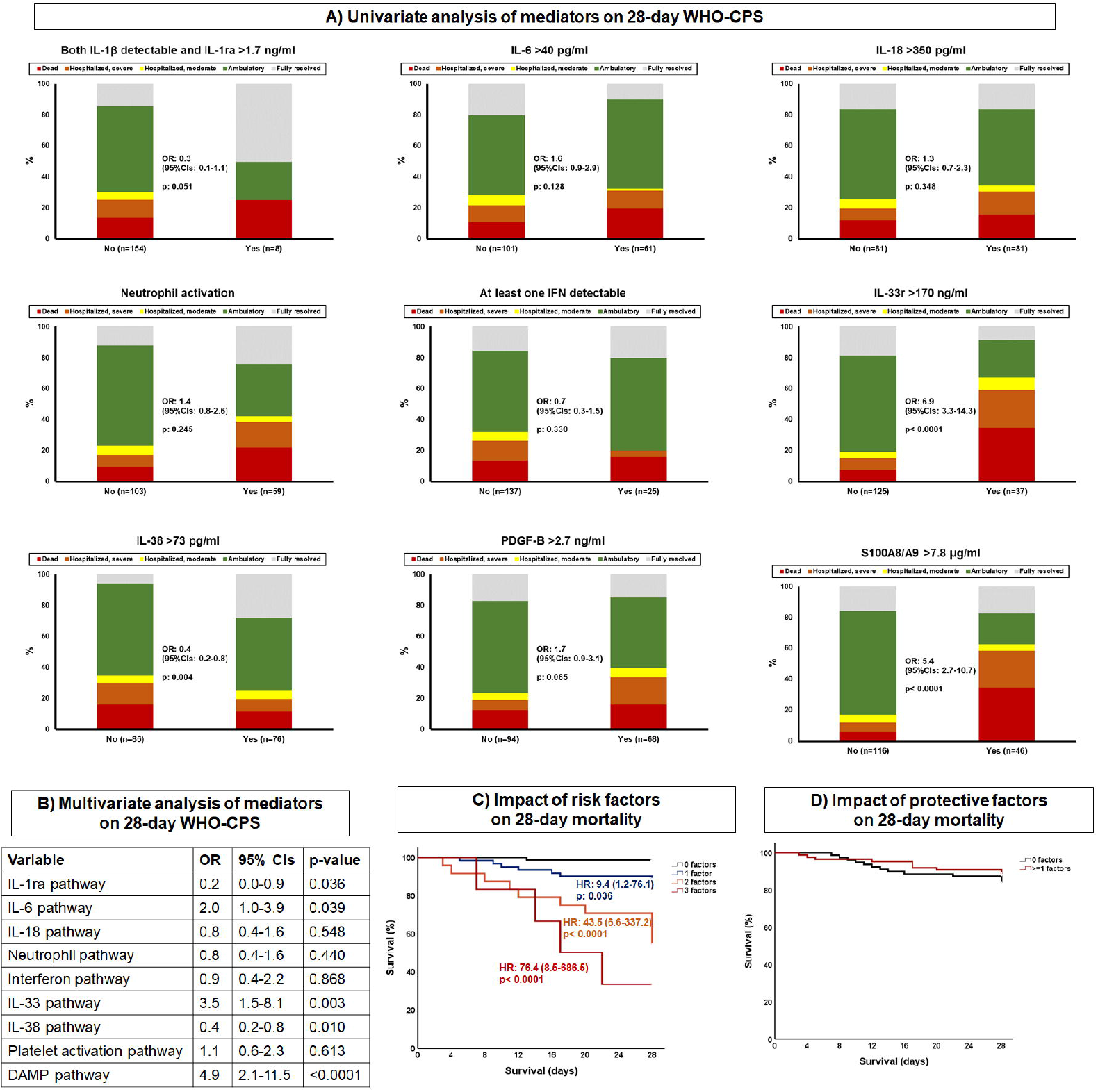
Main drivers of 28-day outcome. The analysis involves patients with COVID-19 hospitalized with moderate COVID-19, severe COVID-19 and hospitalized with ARDS necessitating mechanical ventilation **(A)**. Univariate ordinal regression analysis of factors associated with patterns of activation or inhibition of the inflammatory response. Factors are analyzed by ordinal regression analysis. The principles for the selection of variables used to classify pathways are provided in Supplementary Figure 1. **(B)**. Multivariate ordinal regression analysis of pathways of the inflammatory response **(C)**. Impact of the three factors associated with high-risk for worse outcome coming from the multivariate ordinal regression analysis are analyzed by Cox regression analysis for their impart on 28-day mortality. **(D)**. Impact of the two factors associated with low-risk for worse outcome coming from the multivariate ordinal regression analysis are analyzed by Cox regression analysis for their impart on 28-day mortality. Abbreviations: ARDS=acute respiratory distress syndrome; CI=confidence interval; IL=interleukin; n=number of patients; OR=odds ratio; PDGF=platelet derived growth factor

Multivariate ordinal regression analysis showed that the pathways of IL-6, of IL-33 and of calprotectin were associated with allocation into more severe states of the WHO-CPS after 28 days (Figure 6B). On the contrary, the pathways of IL-1ra and of IL-38 were associated with allocation into less severe states of the WHO-CPS after 28 days. The pathways of IL-6, IL-33 and calprotectin were independent drivers for mortality acting synergistically. On the contrary, the pathways of IL-1ra and IL-38 did not affect survival (Figures 6C and 6D).

### Second stage: validation of the role of S100A8/A9 (calprotectin) for progression into ARDS necessitating MV

The SAVE-MORE trial enrolled patients who were not in either ARDS and MV categories^12^. Among participants allocated to placebo treatment, samples collected serially from patients who progressed into ARDS and MV were analyzed. This was done since patients’s follow-up under placebo are considered to represent the progression of COVID-19 under the current standard-of-care management. At baseline, circulating levels of calprotectin and of PDGF-B were similar among patients who eventually progressed into ARDS and MV and among those who did not progress into ARDS and MV (Figures 7A and 7B). On day 4, there was a trend for increase of calprotectin from baseline among patients who developed ARDS and necessitated MV. This trend became a largely significant difference on day 7 (Figure 7C). No similar changes were found for PDGF-B (Figure 7D). Following ROC curve analysis, it was found that any one-fold or more increase of calprotectin from baseline was associated with the development of ARDS and MV. This association was proven to be independent of disease severity and of dexamethasone treatment after multivariate forward step-wise Cox regression analysis (Figure 7E).

**Figure 7.**
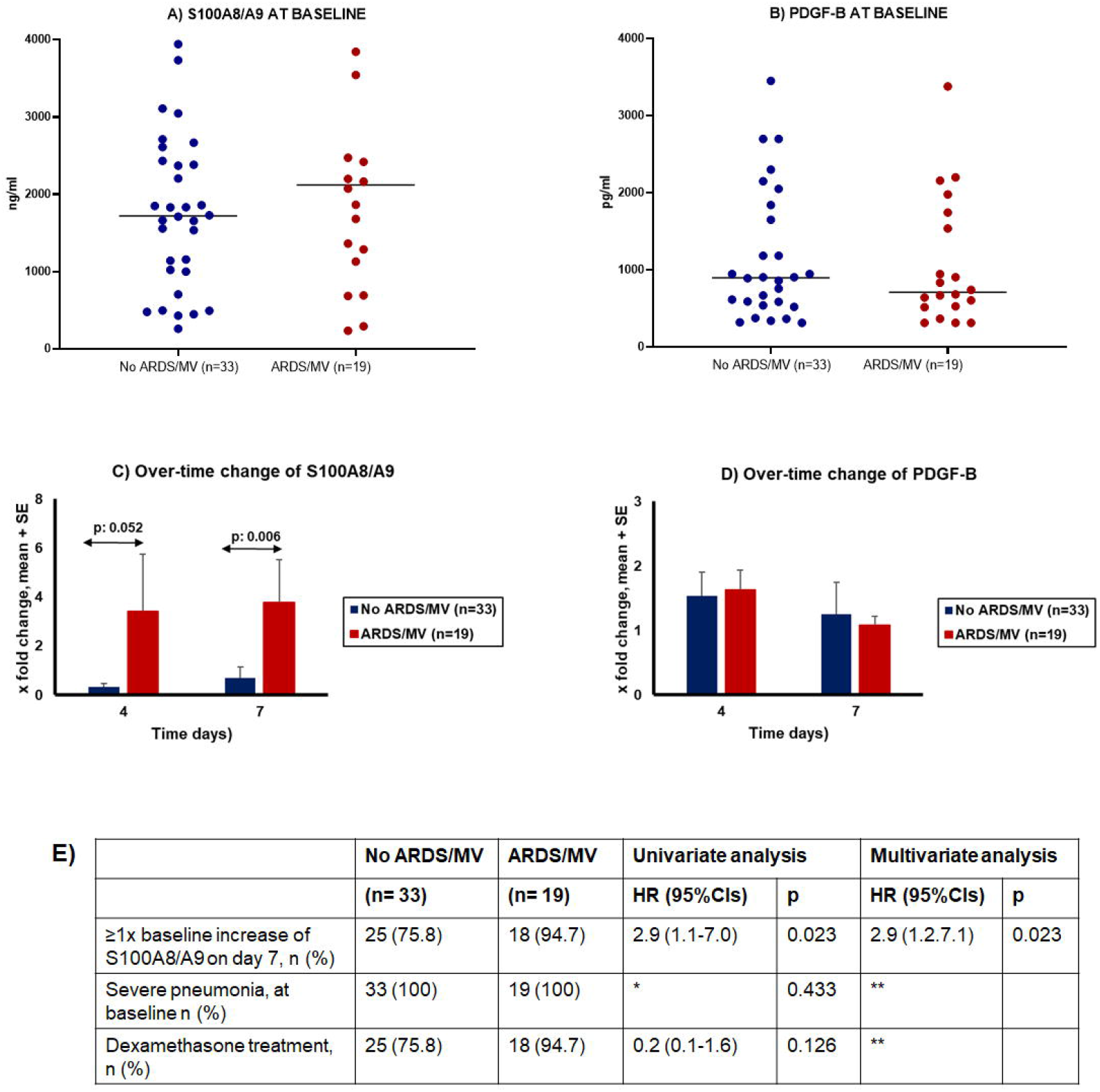
S100A8/A9 (calprotectin) as an independent variable of progression into acute respiratory distress syndrome (ARDS) necessitating mechanical ventilation (MV). The analysis involves serial measurement among 52 patients with severe COVID-19 participating in the study SAVE-MORE. Only statistically significant differences are shown. **(A)**. Circulating concentrations of calprotectin at baseline **(B)**. Circulating concentrations of PDGF-B at baseline **(C)**. Changes of circulating concentrations of calprotectin on days 4 and 7 of follow-up from baseline. **(D)**. Changes of circulating concentrations of PDGF-B on days 4 and 7 of follow-up from baseline **(E)**. Univariate and multivariate forward step-wise Cox regression analysis of variables associated with progression into ARDS and MV. ordinal regression analysis of pathways of the inflammatory response Abbreviations: CI=confidence interval; HR: hazard ratio; PDGF=platelet derived growth factor.

## DISCUSSION

Our results highlight for the first time the conditions necessary for patients with COVID-19 to demonstrate the most worrisome features of critical illness with ARDS necessitating MV. The pathways of danger-associated molecular patterns (DAMPs), IL-33 (ST2) and coagulation are driving ARDS and MV while final outcome is dominated by excess of pro-inflammation through the pathways of IL-6, IL-33 and DAMPs and by decreased levels of the anti-inflammatory pathways IL-38 and IL-1ra.

Our finding of elevated serum level S100A8/A9 (calprotectin) is in line with previous results in the serum^13, 14^ and in the bronchoalveolar lavage of COVID-19 patients^15^ showing, as we did, that calprotectin levels are greater among patients with the more severe state. However, our results are the first to demonstrate that COVID-19 is a disease which is driven in severity by calprotectin. It is known that SARS-CoV-2 per se cannot elicit large amounts of inflammatory mediators leading to ARDS and MV^16^. However rapid viral replication in the lungs leads to the destruction of the lung epithelial cells and to the subsequent release of intracellular DAMPs like calprotectin. Our findings suggest that as the disease progresses, and the patient is worsening towards ARDS, released calprotectin increases and this leads the patient to MV. This pathway of DAMPs is acting synergistically with the pathways of IL-6 and IL-33 mediating unfavorable outcomes.

Although PDGF-B was not increased over-time towards progression into ARDS and MV, the increased circulating PDGF-B described herein is interesting given reports of widespread pulmonary microthromboses in lungs from deceased COVID-19 patients^17^.

Our findings of increased levels of IL-1ra and IL-33r are intriguing as serum levels of IL-1β and IL-33 levels are not significantly increased. This apparent oxymoron may be explained by the autocrine mode of action of these cytokines following their production and implies that COVID-19 patients, especially those with severe disease, produce soluble IL-1ra and IL-33r (sST2) in an apparent effort to curtail the effect of IL-1β and IL-33, respectively. Elevated sST2 has also been reported in severe (especially non-surviving), patients with COVID-19^18^. A recent publication reported that analysis of published scRNseq data from bronchoalveolar lavage fluid from patients with mild to severe COVID-19 contained a population of cells that produce IL-33 and correlate with the severity of the disease^19^. Two other publications reported a unique correlation of IL-12p70/IL-33 with disease severity^20^ and increased expression of IL-33 in cultured epithelial cells infected with SARS-CoV-2^21^. Taken together, these findings seem to imply that there may be local synthesis of IL-33 especially in the lungs leading to reactive production of IL-33r (sST2). IL-33 can stimulate resident mast cells to produce impressive amounts of proinflammatory cytokines^22,23^, and the findings of increased IL-33r should be interpreted as counterbalance to the increased IL-33.

Similar to the increased levels of IL-1ra and IL-33r, the anti-inflammatory IL-38 was increased only in the asymptomatic patients as compared to non-infected comparators but decreased significantly in the more severe patient groups. This finding implies that patients cannot produce sufficient IL-38 to counteract the pro-inflammatory cytokines produced during COVID-19. IL-38 belongs to the IL-1 family^24^ and has been reported to have anti-inflammatory activity^25^. IL-38 exists intracellularly as a precursor full-length form and must be cleaved at the N-terminus before it is secreted extracellularly as active forms^26^. We showed that IL-38 can inhibit release of IL-1β from cultured human microglia stimulated by bacterial lipopolysaccharide (LPS)^27^.

At the end, the final outcome of the patient appears to be determined by the excess activation of the pathways of IL-6, IL-33 and DAMPs, while the anti-inflammatory pathways IL-1ra and IL-38 are not sufficient to counterbalance them. A recent paper reported that elevated plasma levels of IL-6, IL-10 and IP-10 anticipated clinical progression of COVID-19 patients^28^. Our results are in line with the positive treatment outcomes with IL-6 receptor antagonists and with anakinra (the recombinant non-glycosylated form of IL-1ra) because they have showed the need to attenuate the exaggerated IL-6 responses and to enhance the weakened IL-1ra responses^29^. Tocilizumab and sarilumab were shown to decrease the number of days under organ dysfunction and 28-day mortality in the platform adaptive trials REMAP-CAP^30^ and RECOVERY^31,^ respectively. In these trials, the two IL-6 receptor antagonists were administered in patients already in critical illness. In the open-label SAVE trial^32^ and in the double-blind, randomized trial SAVE-MORE^12^ anakinra was administered in hospitalized non-critical patients with plasma levels of the biomarker suPAR (soluble urokinase plasminogen activator receptor 6 ng/ml or more); suPAR was used in these studied as a biomarker of an attack of the host by DAMPs.

Our present results suggest that the innate increase in IL-1ra and IL-33r, along with the drop in IL-38, are not sufficient to halt the progression of COVID-19. Attenuating the pathways of IL-6, IL-33 and DAMPs in parallel with enhancing the pathways of IL-1ra and IL-33r, and potentially administration of recombinant IL-38, seem the most promising strategies to combat critical COVID-19.

## Data Availability

All data produced in the present study are available upon reasonable request to the authors

## Acknowledgments

The authors wish to express their gratitude to Bio-Techne (Minneapolis, MN) for providing the reagent kits. We are also very grateful for generous funding from the Hellenic Institute for the Study of Sepsis. Collection of biomaterial and clinical information from patients enrolled in the ESCAPE trial was funded by the Horizon 2020 Grant RISKinCOVID.

## Author Contributions

GK, AS, GP, GA, AR, HM, GND, PP, SS, SM and ZA collected patient data, revised the manuscript and gave permission for the version to be submitted.

TG performed measurements, revised the manuscript and gave permission for the version to be submitted.

EGB supervised the study, analyzed and plotted the results, wrote the manuscript and takes full responsibility of data integrity.

TCT conceived the study, secured the reagents, participated in data analysis and wrote the manuscript.

## METHODS

### Patient cohorts

Studied patients were adults with molecular detection of SARS-CoV-2 and classified into four groups using the WHO classification of COVID-19 severity: (a) asymptomatic; (b) hospitalized with moderate COVID-19; (c) hospitalized for severe COVID-19; and (d) hospitalized with acute respiratory distress syndrome (ARDS) necessitating mechanical ventilation. Ten ml of blood samples were collected the first 24 hours from admission in the emergency department or in the general ward or in the intensive care unit and collected into pyrogen-free tubes (Becton Dickinson, Cockeysville Md). Patients have already participated as comparators receiving usual care in the studies ACHIEVE^33^, SAVE^32^ and ESCAPE^34^ that have been approved by the National Ethics Committee of Greece and by the National Organization for Medicines of Greece (approval numbers 45/20 and IS 36/20 respectively for ACHIEVE; approval numbers 38/20 and IS 28/20 for SAVE; and approval numbers approval 30/20 and IS 021-20 respectively for ESCAPE). Patients were enrolled after written informed consent provided by themselves or legal representatives.

Ten ml of blood were also collected from comparators matched for age and sex with the patients and who were admitted at the emergency departments for unspecified complaints. These comparators were adults without any specific diagnosis of disease as this was certified by a phone call after 30 days. Their inclusion was provisioned in the above studies and they also signed a written informed consent.

In order to provide evidence for the role of the mediators for progression into ARDS necessitating MV, samples collected from 52 patients participating in the SAVE-MORE trial were studied. SAVE-MORE is a double-blind randomized clinical trial where patients were allocated to treatment with placebo or anakinra plus standard-of-care to prevent progression into severe respiratory failure^12^ (approval 161/20 by the National Ethics Committee of Greece; approval 01.02.2021 by the Ethics Committee of the National Institute for Infectious Diseases Lazzaro Spallanzani, IRCCS in Rome; EudraCT number, 2020-005828-11; ClinicalTrials.gov NCT04680949). The studied samples were coming from patients with severe disease in need of treatment with oxygen and who were allocated to the placebo group so that comparative levels of the mediators at serial time points may dictate their role towards development into ARDS and MV. For this aim only patients from the placebo group not being under modulation with biologicals with all three time periods of sampling available (baseline, day 4 and day 7) were studied; 19 patients progressed into ARDS and MV the first 28 days; another 33 patients matched for age, gender and comorbidities who did not progress into ARDS and MV were studied as comparators.

### Laboratory assays

All blood samples were processed immediately and the serum was stored in - 80°C until assayed. Levels of biomarkers were quantified by using commercially available kits of enzyme immunosorbent assays from Bio-Techne (R&D Systems, Minneapolis, MN) according to the manufacturer’s instructions. The lower limits of detection were: 16 pg/ml for tumor necrosis factor-alpha (TNFα); 8 pg/ml for interleukin (IL)-1β; 31 pg/ml for IL-1ra (receptor antagonist); 40 pg/ml for IL-6; 31 pg/ml for IL-8 and IL-10; 8 pg/ml for IL-17; 62 pg/ml for IL-18; 31 pg/ml for IL-33r (receptor); 62 pg/ml for IL-38; 156 pg/ml for interferon (IFN)-alpha; 78 pg/ml for IFNβ; 156 pg/ml for IFNγ; 80 pg/ml for platelet activation factor (PAF); 313 p/ml for platelet-derived growth factor (PDGF)-A and PDGF-B; 61 pg/ml for S100A8/A9; 31 pg/ml for matrix metallpeptidase (MMP)-9; and 46 pg/ml for S100B.

### Study endpoints

The study primary endpoint was to define a set of cytokines and inflammatory mediators that are independently driving the progression into critical ARDS necessitating mechanical ventilation.

The study secondary endpoints were to define a set of cytokines and inflammatory mediators which are independently associated with the outcome of COVID-19 after 28 days as this is expressed both by the strata of the WHO Clinical Progression Scale (CPS) and by mortality.

### Statistical analysis

Concentrations of biomarkers between groups of patients were compared using the Mann-Whitney U non-parametric test following Bonferroni correction for multiple testing. Receiver operator characteristics (ROC) curve analysis was done for each of the measured mediators to define a cut-off that can significantly discriminate patients with ARDS necessitating MV; the areas under the curve of the ROC (AUC), 95% confidence intervals (CIs) and p-values were determined. For variables which were significant, the cut-off concentration providing the best trade-off for sensitivity and specificity was determined by the Youden index. In order to define the variables which were drivers of ARDS necessitating MV forward step-wise logistic regression analysis was done. ARDS necessitating MV entered the equation as dependent variable and all significant mediators at the pre-defined cut-offs as independent variables.

Mediators were divided into a pathway-like approach as follows: a pathway-like division of biomarkers. To this end, nine pathway were defined as follows: a) IL-1ra activation using IL-1β and IL-1ra; b) IL-6 pathway using IL-6; c) IL-18 pathway using IL-18; d) neutrophil activation using IL-8, MMP-9 and IL-17; e) interferon pathway using the three measured IFNs; f) IL-33 pathway using IL-33 and IL-33r; g) IL-38 pathway using IL-38; h) platelet activation pathway using PAF, PDGF-A and PDGF-B; and i) calprotectin/alarmin pathway using S100A8/A9. The pathway was defined in a hierarchical order using as first step the cut-offs of ROC curve analysis in case of mediators being significant; and as second step the median levels of the entire cohort (Figure 5). Patients were sub-grouped for each of the nine pathways into those belonging or not-belonging to this pathway of activation (Yes/No). Comparisons of the WHO-CPS strata at each sub-group were done by univariate ordinal regression analysis followed by multivariate ordinal regression analysis. 28-day mortality was compared between subgroups by Cox forward step-wise multiple regression analysis. 28-day mortality was the dependent variable and variables being significant for WHO-CPS were the independent variables.

The fold-change of mediators shown from the first stage of analysis to be associated with ARDS and MV on days 4 and 7 from baseline was calculated and compared by the Mann-Whitney U test. The cut-off of fold-change that was associated with ARDS and MV was defined by the area under the ROC curve using the Youden index. The value of this cut-off was further validated by forward step-wise Cox regression analysis. Progression into ARDS and MV was the dependent variables; the defined cut-offs, baseline severity and dexamethasone treatment were the independent variables.

## Data Availability

Requests for deidentified patient data by researchers with proposed use of the data can be made to corresponding author with specific data needs, analysis plans and dissemination plans.

## References

1. Tai, W. et al. Characterization of the receptor-binding domain (RBD) of 2019 novel coronavirus: implication for development of RBD protein as a viral attachment inhibitor and vaccine. Cell. Mol. Immunol. 17, 613–620 (2020).

2. Canna, S.W. & Cron, R.Q. Highways to hell: Mechanism-based management of cytokine storm syndromes. J. Allergy Clin. Immunol. 146, 949–959 (2020)

3. Moore, J.B. & June, C.H. Cytokine release syndrome in severe COVID-19. Science 368, 473–474 (2020)

4. Giamarellos-Bourboulis, E.J. et al. Complex immune dysregulation in COVID-19 patients with severe respiratory failure. Cell Host Microbe 27, 992–1000 (2020)

5. Chen, G. et al. Clinical and immunological features of severe and moderate coronavirus disease 2019. J. Clin. Invest. 130, 2620–2629 (2020)

6. Brodin, P. Immune determinants of COVID-19 disease presentation and severity. Nat. Med. 27, 28–33 (2021)

7. Herold, T, et al. Elevated levels of IL-6 and CRP predict the need for mechanical ventilation in COVID-19. J. Allergy Clin. Immunol. 146, 128–136 (2020)

8. Han, H., et al. Profiling serum cytokines in COVID-19 patients reveals IL-6 and IL-10 are disease severity predictors. Emerg. Microbes Infect. 9, 1123–1130 (2020)

9. Mazzoni, A., et al. Impaired immune cell cytotoxicity in severe COVID-19 is IL-6 dependent. J. Clin. Invest. 130, 4694–4703 (2020)

10. Conti, P. et al. Coronavirus-19 (SARS-CoV-2) induces acute severe lung inflammation via IL-1 causing cytokine storm in COVID-19: a promising inhibitory strategy. J. Biol. Regul. Homeost. Agents 34, 1971–1975 (2020)

11. Mahler, M., Meroni, P.L., Infantino, M., Buhler, K.A. & Fritzler, M.J. Circulating calprotectin as a biomarker of COVID-19 severity. Expert Rev. Clin. Immunol. 17, 431–443 (2021)

12. Kyriazopoulou, E. et al. Early treatment of COVID-19 with anakinra guided by soluble urokinase plasminogen activator receptor: a double-blind, randomized controlled phase 3 trial. Nat. Med. 27, 1752–1760 (2021)

13. Chen, L. et al. Elevated serum levels of S100A8/A9 and HMGB1 at hospital admission are correlated with inferior clinical outcomes in COVID-19 patients. Cell. Mol. Immunol. 17, 992–994 (2020)

14. Bauer, W., et al. Outcome prediction by serum calprotectin in patients with COVID-19 in the emergency department. J. Infect. 82, 84–123 (2021).

15. Huang, W. et al. The inflammatory factors associated with disease severity to predict COVID-19 progression. J. Immunol. 206, 1597–1608 (2021)

16. Paludan, S.R. & Mogensen, T.H. Innate immunological pathways in COVID-19 pathogenesis. Science Immunol. 7, eabm5505 (2022).

17. Ackermann, M. et al. Pulmonary vascular endothelialitis, thrombosis, and angiogenesis in Covid-19. N. Engl. J. Med. 383, 120–128 (2020)

18. Zeng, Z. et al. Serum-soluble ST2 as a novel biomarker reflecting inflammatory status and illness severity in patients with COVID-19. Biomark. Med. 14, 1619–1629 (2020)

19. Stanczak, M.A. et al. IL-33 expression in response to SARS-CoV-2 correlates with seropositivity in COVID-19 convalescent individuals. Nat. Commun. 12, 2133 (2021)

20. Munitz, A. et al. Rapid seroconversion and persistent functional IgG antibodies in severe COVID-19 patients correlates with an IL-12p70 and IL-33 signature. Sci. Rep. 11, 3461 (2021)

21. Liang, Y., Ge, Y. & Sun, J. IL-33 in COVID-19: friend or foe? Cell. Mol. Immunol. 18, 1602–1604 (2021)

22. Taracanova, A. et al. SP and IL-33 together markedly enhance TNF synthesis and secretion from human mast cells mediated by the interaction of their receptors. Proc Natl Acad Sci U S A 114, E4002–E4009 (2017)

23. Taracanova, A., Tsilioni, I., Conti, P., Norwitz, E.R., Leeman, S.E. & Theoharides, T.C. Substance P and IL-33 administered together stimulate a marked secretion of IL-1beta from human mast cells, inhibited by methoxyluteolin. Proc Natl Acad Sci U S A 115, E9381–E9390 (2018)

24. Mantovani, A., Dinarello, C.A., Molgora, M. & Garlanda, C. Interleukin-1 and related cytokines in the regulation of inflammation and immunity. Immunity 50, 778–795 (2019)

25. van de Veerdonk, F.L., de Graaf, D.M., Joosten, L.A. & Dinarello, C.A. Biology of IL-38 and its role in disease. Immunol. Rev. 281, 191–196 (2018)

26. Kwak, A., Lee, Y., Kim, H. & Kim, S. Intracellular interleukin (IL)-1 family cytokine processing enzyme. Arch. Pharm. Res. 39, 1556–1564 (2016)

27. Tsilioni, I., Pantazopoulos, H., Conti, P., Leeman, S.E. & Theoharides, T.C. IL-38 inhibits microglial inflammatory mediators and is decreased in amygdala of children with autism spectrum disorder. Proc Natl Acad Sci U S A 117, 16475–16480 (2020)

28. Laing AG, et al. A dynamic COVID-19 immune signature includes associations with poor prognosis. Nat Med. 26, 1623–1635 (2021).

29. Dalekos, G.N., et al. Lessons from pathophysiology: use of individualized combination treatments with immune interventional agents to tackle severe respiratory failure in patients with COVID-19. Eur. J. Intern. Med. 88, 52–62 (2021)

30. Gordon, A.C. et al. Interleukin-6 receptor antagonists in critically ill patients with Covid-19. N. Engl. J. Med. 384, 1491–1502 (2021)

31. Abani, O. et al. Tocilizumab in patients admitted to hospital with COVID-19 (RECOVERY): a randomised, controlled, open-label, platform trial. Lancet 397, 1637–1645 (2021)

32. Kyriazopoulou, E. et al. An open label trial of anakinra to prevent respiratory failure in COVID-19. eLife 10, e66125 (2021)

33. Tsiakos, K. et al. Early start of clarithromycin is associated with better outcome in COVID-19 of moderate severity: the ACHIEVE open-label single-arm trial. Infect. Dis. Ther. 10, 2333–2351 (2021)

34. Karakike, E. et al. ESCAPE: An open-label trial of personalized immunotherapy in critically ill COVID-19 patients. J. Innate Immun. doi: 10.1159/000519090 (2021)

