## Supplement for "Calprotectin and inflammation-associated serum biomarkers determine critical illness in COVID-19"

**Theoharis C. Theoharides^12,13,14,15^***

*equal contribution
^1^Intensive Care Unit, Korgialeneion-Benakeion Athens General Hospital, 115 27 Athens, Greece;

^2^4^th^ Department of Internal Medicine, National and Kapodistrian University of Athens, Medial School, 124 62 Athens, Greece;

^2^3^rd^ Department of Internal Medicine, National and Kapodistrian University of Athens, Medical School, 115 27 Athens, Greece;

^4^1^st^ Department of Internal Medicine, G. Gennimatas General Hospital of Athens, 115 27 Athens, Greece;

^5^2^nd^ Department of Pulmonary Medicine, Sotiria General Hospital of Chest Diseases, 115 27 Athens, Greece;

^6^1st Department of Internal Medicine, University of Ioannina, Medical School, 455 00 Ioannina, Greece;

^7^Department of Medicine and Research Laboratory of Internal Medicine, National Expertise Center of Greece in Autoimmune Liver Diseases, General University Hospital of Larissa, 41110 Larissa, Greece;

^8^2^nd^ Department of Internal Medicine, Democritus University of Thrace, Medical School, 681 00 Alexandroupolis, Greece;

^9^1^st^ Department of Internal Medicine, Thriasio General Hospital of Eleusis, 196 00 Magoula, Greece;

^10^1st Department of Internal Medicine, Aristotle University of Thessaloniki, Medical School, 546 21 Thessaloniki, Greece;

^11^2nd Department of Internal Medicine, Thriasio General Hospital of Eleusis, 196 00 Magoula, Greece;

^12^Laboratory of Molecular Immunopharmacology and Drug Discovery, Department of Immunology, Tufts University School of Medicine, Boston. MA 02111, USA

^13^School of Graduate Biomedical Sciences, Tufts University School of Medicine, Boston, MA 02111, USA

^14^Department of Internal Medicine, Tufts University School of Medicine and Tufts Medical Center, Boston, MA 02111, USA

^15^Institute of Neuro-Immune Medicine, Nova Southeastern University, Clearwater , FL 33759, USA

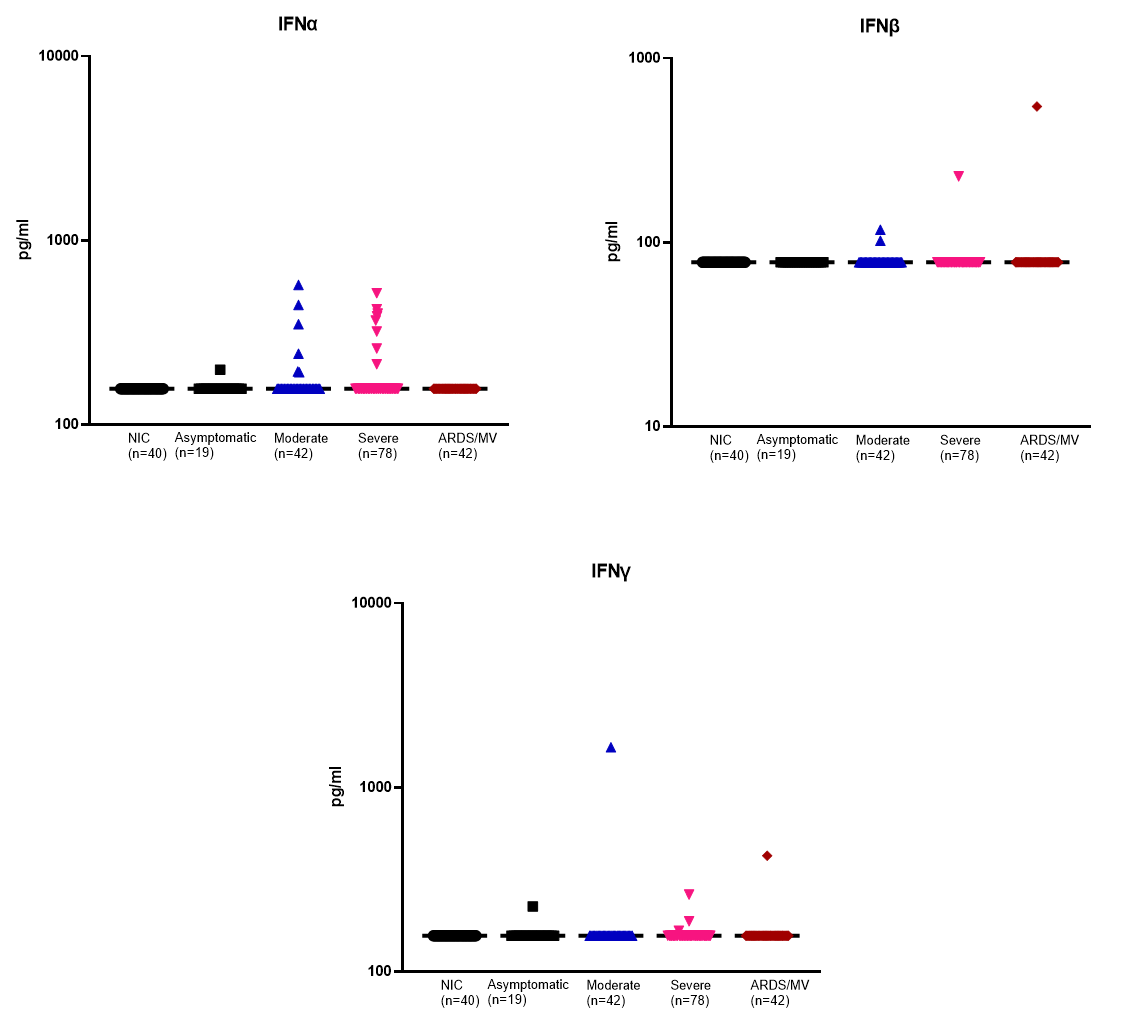

**Supplementary Figure 1. Concentrations of interferons.**

Dot plots with horizontal lines indicating the median of each group. The numbers of subjects evaluated are listed in the parentheses. Double arrowhead lines indicate comparisons between groups. Only statistically significant differences are indicated as follows: *p <0.05; ** p <0.01; ***p< 0.001; ****p< 0.0001. Abbreviations: ARDS= acute respiratory distress syndrome; IFN= interferon; MV= mechanical ventilation; n=number of patients; NIC= non-infected comparators
